# Prevalence of Coronary Atherosclerosis in Master Female Endurance Athletes

**DOI:** 10.1101/2023.11.21.23298867

**Authors:** Efstathios Papatheodorou, Vincent L. Aengevaeren, Thijs M.H. Eijsvogels, Khaled Al Fakih, Rebecca Kathryn Hughes, Ahmed Merghani, Christine K. Kissel, Saad Fyyaz, Athanasios Bakalakos, Mathew G. Wilson, Damini Dey, Gherardo Finocchiaro, Gemma Parry-Williams, Camilla Torlasco, Michael Papadakis, James C. Moon, Sanjay Sharma

**Affiliations:** Cardiology Clinical and Academic Group. St Georges, University of London, and St George’s University Hospital NHS Foundation Trust (UK); Department of Medical Biosciences, Exercise Physiology Research Group. Radboud University Medical Centre. Nijmegen, the Netherlands; University Hospital Lewisham, London (UK); The Barts Heart Centre, University College London (UK); Institute of Sport and Exercise Health (ISEH), University College London, London, UK; HCA Healthcare Research Institute, London, UK; Cedars-Sinai Medical Centre, Biomedical Imaging Research Institute, Los Angeles, CA (USA)

## Abstract

**BACKGROUND:** Studies in ostensibly healthy male master athletes have revealed a greater prevalence of coronary artery calcification (CAC) and coronary plaques compared with relatively sedentary counterparts. In contrast, data relating to potentially adverse coronary remodelling in female master athletes is sparse and conflicting. We investigated the prevalence of coronary atherosclerosis in a cohort of predominantly post-menopausal female master athletes with a low atherosclerotic risk profile.

**METHODS:** 196 female athletes with a mean age of 55±7 years-old and median exercise duration of 33 years (25-39) and 59 relatively sedentary females of similar age underwent cardiovascular investigations including a coronary computed tomogram angiography with assessment of CAC scores, coronary plaques, and pericoronary adipose tissue attenuation. 70% of the athletes and 68% of the control group were post-menopausal.

**RESULTS:** Athletes and controls had a similarly low Framingham 10-year risk (1.49% versus 2.1%; P=0.68), but body mass index and blood pressure were lower and HDL-C was higher in the athletes. The prevalence of CAC score >0 Agatston units (AU) was low and did not differ between athletes and controls (21% *versus* 32%;P=0.073). Female athletes had a lower prevalence of a CAC score >50^th^ centile (19% *versus* 32%;P<0.03) and >75^th^ centile (14% *versus* 25%;P=0.045) for age compared with controls, but the prevalence of a CAC score >100 AU did not differ between the groups (3.6% *versus* 8.5%;P=0.12). There were also no differences between the groups in the prevalence of individuals with coronary plaques (21% *versus* 32%;P=0.09), total plaque volume (16 mm^3^ [IQR 3-56] *versus* 49 [5-142] mm^3^;P=0.08), or plaque burden (10.8% [2.8-21] *versus* 15.4% [4.6-28];P=0.46). Coronary plaques were predominantly calcified in both athletes and controls (80% *versus* 63%;P=0.08). Age, blood pressure and HDL-C were independent predictors for a CAC score >0 AU among athletes.

**CONCLUSIONS:** In contrast with previous studies in male master athletes, lifelong exercise in female counterparts does not appear to be associated with increased CAC score, coronary plaque burden or any qualitative differences in coronary plaque compared with relatively sedentary healthy counterparts. Coronary atherosclerosis in master female athletes is mainly driven by traditional risk factors.

## Introduction

The benefits of exercise for curbing risk factors for atherosclerosis and reducing overall cardiovascular morbidity and mortality are widely recognized^1,2^, however, several studies have shown an increased burden of coronary atherosclerosis in male master athletes^3–6^. Whereas original studies reported increased coronary artery calcification (CAC) in male marathon runners with a relatively high prevalence of atherosclerotic risk factors^4^, recent studies have shown similar findings in male master athletes with a low atherosclerotic risk profile ^3,5,6^. Increasing age, traditional risk factors and volume of exercise are major determinants of high CAC scores in males although coronary inflammation, dietary factors, blood pressure responses to exercise and serum parathyroid hormone concentrations have also been postulated^7–9^. Over the past couple of decades, there has been an explosive surge in the number of middle-aged and older women participating and excelling in mass endurance sporting events and competitive races, however there are few studies examining the effect of lifelong exercise on the coronary vasculature in women. Existing reports are conflicting^6,10–12^ and limited by the small number of participants, variation in the prevalence of atherosclerotic risk factors between the comparison groups and inclusion of mainly premenopausal or perimenopausal women who may be protected by the anti-atherogenic effects of estrogen.

We investigated the prevalence, magnitude and determinants of CAC score and coronary plaque in predominantly post-menopausal female master athletes with a low atherosclerotic risk profile.

## METHODS

### Subjects

Female master athletes were defined as athletes aged ≥40 years-old who had exercised systematically and continuously for ≥10 years and had competed in ≥10 endurance events. Athletes were recruited by contacting governing sporting bodies for master athletes including the British Master Athletics Federation, British Master Swimming Association, Master Rowing, British triathlon and several endurance sports clubs in London, United Kingdom. Healthy controls were recruited through advertisements in 3 London National Heart Service Trusts and advertisements in community centers. Controls were of similar age and allowed to engage in up to 3 hours of moderate intensity physical activity and recreational exercise per week. Athletes or controls with a history of coronary artery disease (CAD), hypertension, diabetes mellitus, a Framingham 10-year risk score >10%, a family history of premature CAD (<65 years in female relatives or <55 years in male relatives), current or previous history of smoking, and contraindications to coronary computed tomography angiography (CCTA), or cardiovascular magnetic resonance (CMR) were excluded. Between 02/2017 and 09/2018 196 athletes and 59 sedentary controls underwent a battery of investigations including a health questionnaire, blood tests, ECG, echocardiogram, cardiopulmonary exercise test (CPET), coronary computed tomography angiogram (CCTA) and cardiovascular magnetic resonance (CMR) scan. Participants attended on 3 separate days to complete these investigations. The first attendance included a health questionnaire, blood tests and CPET. The second and third attendances involved a CCTA and CMR respectively. The time span for completing all these investigations per participant was 4-6 weeks.

### Health questionnaire

Participants completed a baseline questionnaire (supplement 1) enquiring about demographics, cardiac symptoms, medical history, smoking history, family history of cardiac disease, menstrual history including age at menopause, if applicable, and exercise history. Menopause was considered if a woman had not menstruated for ≥12 months.

### Exercise history and intensity

The exercise history focused on the duration and intensity of exercise, including total number of years of exercise, sports discipline(s), total number of competitions in sporting events, best times achieved in competitions, and major achievements. The intensity of exercise was assessed through the International Physical Activity Questionnaire (IPAQ) which is a widely used, standardized, and validated questionnaire^13,14^ (supplement 2). The weekly dose of exercise was estimated by calculating the metabolic equivalent of task (MET) minutes per week from the information ascertained by the IPAQ.

### Blood investigations

Blood samples were analyzed for serum lipid profile, blood glucose and renal function. A separate blood sample was also taken 15 minutes after the completion of the cardiopulmonary exercise test for assessment of parathyroid hormone (PTH) concentrations, with values > 6.9 pmol/L to be considered as increased (laboratory reference range 1.1 – 6.9 pmol/L).

### Cardiopulmonary exercise test

Cardio-pulmonary exercise testing was performed in an upright position using a COSMED E100w cycle ergometer (Rome, Italy) with an incremental ramp protocol of 20-25 Watts/min for athletes and 10-15 Watts/min for healthy controls and simultaneous continuous ECG recording. Subjects were encouraged to exercise to the point of exhaustion. Breath-by-breath gas exchange analysis was performed using a dedicated COSMED Quark CPEX metabolic cart (Rome, Italy) and standard cardiometabolic indices were obtained according to published guidelines^15^. Blood pressure was manually obtained at rest and at 2-minute intervals during exercise. Exercise induced hypertension was defined as systolic blood pressure of ≥190mmHg during exercise^22,23^.

### Cardiac Investigations

#### Coronary computed tomography angiography (CCTA)

CCTA was performed using a 64 slice LightSpeed VCT XTe GE scanner (GE Healthcare) and prospective gating, using a commercially available protocol (SnapShot Pulse, GE healthcare) and the following scanning parameters: slice acquisition 64 x 0.625 mm, smallest X-ray window, Z-coverage value of 20 mm with an increment of 20 mm, gantry rotation time of 350 ms. Some participants received intravenously Metoprolol (5-20mg) to achieve a heart rate ≤60 bpm. All volunteers received two 400 mcg doses of sublingual Glyceryl trinitrate. The coronary circulation was visualized by injecting 100 ml of intravenous Ivoersol (Optiray 350 mg I/ml, Covidien UK, Hampshire, UK) at a flow rate of 5 ml/s followed by 100 ml of saline solution into an antecubital vein via an 18-gauge peripheral venous catheter. All images were transferred to an external workstation (Autoplaque, v2.18.08.23, Mount Sinai, USA) for analysis. Plaque quantification analysis was performed in 35 athletes and 10 controls. A semi-automated 3D quantification and characterization of segments with plaque on CCTA was used to define the plaque composition, total plaque volume and total plaque burden as validated previously^16,17^. Plaque prevalence was defined as the number of individuals with plaques. Plaque burden was defined as the respective plaque volume/vessel volume. Plaque morphology was sub-divided into calcified, mixed morphology or non-calcified plaque and plaque composition was defined as plaque morphology/total plaque volume. Pathological CAD was defined as the presence of ≥1 of the following (a) CAC score >100 Agatston Units (AU), (b) CAC score >75% for age, or (c) vessel stenosis>50%.

### Pericoronary Adipose Tissue (PCAT)

PCAT CT attenuation was used to assess coronary inflammation based on previously described and validated methodology^18–20^. PCAT was defined as the adipose tissue within a radial distance from the outer vessel wall equal to the diameter of the vessel. Quantification was based on the attenuation histogram of perivascular fat within the range –190 Hounsfield Units (HU) to –30 HU. The proximal right coronary artery (RCA) was used for PCAT analysis. The proximal 10-50mm of the RCA were analyzed excluding the first 10mm. PCAT CT attenuation derived values were normalized for the kilovoltage peak (KVP) potential. The mean PCAT CT attenuation was determined in a 1 mm cylinder within a radial distance from the outer vessel wall equal to the average diameter of the right coronary artery (FAI_RCA_). FAI analysis was performed in 151 female athletes and 36 controls.

### Cardiovascular Magnetic Resonance (CMR) Scan

Participants underwent CMR using a 1.5T magnet (Aera, Siemens Medical Solutions) as described previously^6^. Chamber volumes, myocardial mass, ventricular function, and aortic dimensions were assessed by cine steady-state free precession sequences^21^. Delayed contrast enhancement images were obtained 15 min after intravenous bolus injection of gadolinium-diethylene triamine pentaacetic acid (Gd-DTPA) (0.15 mmol/kg, Magnevist, Bayer Healthcare Pharmaceuticals, New Jersey, USA) to identify regional fibrosis. Inversion times were adjusted to null normal myocardium and late gadolinium enhancement (LGE) images were phase swapped to exclude artefacts.

### Statistical Analysis

Data are expressed as n (%), mean ± standard deviation (SD) for normally distributed continuous variables, or as median plus 25^th^-75^th^ interquartile percentiles for continuous variables that are not normally distributed. Statistical analyses were performed using Stata/IC version 15.1. Continuous variables were tested for normality using a Shapiro-Wilk test. Chi-squared and Fischer’s exact tests were used for the comparison of categorical variables while unpaired t-tests and Mann-Whitney U tests were used for normally and non-normally distributed variables respectively.

Among master athletes, univariate logistic regression was performed to identify statistically significant variables associated with CAC score >0 and FAI. We decided *a priori* to include age, years of exercise, competitions completed, MET mins of exercise per week, menopausal status, resting systolic blood pressure (SBP), maximal SBP during exercise, total cholesterol, HDL cholesterol, parathyroid hormone (PTH) levels, left ventricular mass, and peak VO_2_. Coronary inflammation (FAI_RCA_) was subsequently added to the CAC score >0 model. A backward stepwise multivariable logistic regression model was constructed retaining all variables with associated with CAC score >0 or FAI_RCA_ (P<0.10) in the univariable analyses. A P-value <0.05 was considered statistically significant.

## RESULTS

### Demographics and atherosclerotic risk profile

A total of 196 master athletes and 59 controls fulfilled criteria for participation. Athletes and controls were of similar age and had a similarly low mean Framingham risk and post-exercise PTH concentrations. There were no significant differences between athletes and controls with respect to the proportion of post-menopausal women. Controls were heavier than athletes, which translated to a modestly greater body surface area. Controls also revealed higher systolic and diastolic blood pressure readings, whereas athletes had a slightly higher mean total and HDL cholesterol compared with controls. Athletes revealed higher peak oxygen consumption than controls and almost a quarter of the athletes showed an exaggerated blood pressure during exercise (SBP>190 mm Hg at peak exercise) compared with 10% of the control group (p<0.001) (**Table 1**).

**Table 1.** Baseline demographics, atherosclerotic risk profile characteristics, exercise history and imaging characteristics of female master athletes and sedentary controls.

|  | **Female Athletes**<br>**N=196** | **Controls**<br>**N=59** | **p value** |
| --- | --- | --- | --- |
| Age (years) | 54.8 ± 7.4 | 54 ± 6.4 | 0.99 |
| Height (cm) | 166.4 ± 6.4 | 163.7 ± 7.4 | **<0.01** |
| Weight (kg) | 59 ± 8.1 | 66 ± 6 | **<0.01** |
| BMI (kg/m<sup>2</sup>) | 21.4 ± 4.1 | 24 ± 5.5 | **<0.01** |
| White Caucasian n (%) | 180 (92) | 55 (93) | 0.92 |
| ***Risk factors for CAD*** |  |  |  |
| Systolic BP (mm Hg) | 115 ± 6.6 | 124 ± 6.9 | **<0.01** |
| Diastolic BP (mm Hg) | 74.3 ± 7.6 | 78.3 ± 7.8 | **<0.01** |
| Total Cholesterol (mmol/L) | 5.0 ± 0.5 | 4.8 ± 0.6 | **<0.01** |
| HDL-C (mmol/L) | 2.4 ± 0.3 | 1.9 ± 0.3 | **<0.01** |
| Menopausal n (%) | 139 (71) | 40 (68) | 0.85 |
| Framingham 10-year risk score | 1.5 ± 0.6 | 2.1 ± 0.7 | 0.09 |
| ***Exercise History*** |  |  |  |
| Years of Exercise | 33 (25 - 39) | 2 (0 - 1) | **<0.01** |
| Hours of exercise per week | 8 (6 - 10) | 0 (0 - 1) | **<0.01** |
| Days of exercise per week | 6 (4 - 6) | 0 (0 - 2) | **<0.01** |
| Total Competitions | 90 (15 - 134) | 0 (0 - 0) | **<0.01** |
| METS minutes per week | 7225 (4784 – 10647) | 233 (120-919) | **<0.01** |
| Vigorous METS minutes per week | 2880 (2160 – 4800) | 0 (0 - 280) | **<0.01** |
| ***CPET and CMR parameters*** |  |  |  |
| Peak VO<sub>2</sub> (ml/min/kg) | 42 ± 4.6 | 25 ± 7.3 | **<0.01** |
| PeakVO<sub>2</sub> predicted (%) | 163.5 ± 24.6 | 104.8 ± 19.9 | **<0.01** |
| Exercise induced HTN n (%) | 48 (25%) | 6 (10%) | **<0.01** |
| Post exertional PTH levels (pmol/l) | 7.84 ± 2.5 | 8.3 ± 2.4 | 0.40 |
| Raised post-exertional PTH n (%) | 114 (58) | 35 (57) | 0.91 |
| LVEDV (ml) | 137 ± 23 | 121 ± 21 | **<0.01** |
| LVEDV/BSA (ml/mm<sup>2</sup>) | 82 ± 12 | 69 ± 8 | **<0.01** |
| RVEDV (ml) | 132 ± 26 | 124 ± 27 | **<0.01** |
| RVEDV/BSA (ml/mm<sup>2</sup>) | 80 ± 14 | 69 ± 10 | **<0.01** |
| Left Ventricular Mass (g) | 129.5 ± 32 | 112 ± 25.5 | **<0.01** |
| RV insertion point fibrosis n (%) | 62 (32) | 5 (8) | **<0.01** |
| Ischemic LGE | 2 (1%) | 0 (0%) | 0.54 |

BP=Blood Pressure; CAD=Coronary Artery Disease; CMR=Cardiovascular magnetic
resonance; CPET=Cardiopulmonary exercise test; HDL-C=High-Density Lipoprotein
Cholesterol; HTN=Hypertension; LGE= Late Gadolinium Enhancement; LVEDV = LV
End-diastolic Volume; METS=Metabolic Equivalents; PTH=parathyroid hormone;
RVEDV = RV End-diastolic Volume; SD = Standard Deviation; VO2 = Oxygen
Consumption. Values are presented as mean ± SD, n (%), or median (Interquartile

### Exercise history

Among athletes, endurance competitions included marathons (26.2miles; 42.2km), half marathons (13.1miles; 21.1km), 10km running races, endurance cycling races ranging from 41.1 miles to 161.5miles; 66 to 260km), triathlons, rowing regattas, marathons, or river races. Master athletes had been exercising for a median of 33 years and trained for an average of 8 hours per week. Forty-five athletes were UK Master champions, 27 were European Master champions or finalists, 16 were World Master champions, and 2 were former Olympic medalists. Most athletes engaged in running (n=144, 73%), followed by cycling (n=91, 46%), swimming (n=82, 41%) and rowing (n=22, 11%).

### Coronary calcium score

The prevalence of CAC score >0 was low and did not differ between athletes and controls (21% versus 32%: P=0.073) (**Table 2**). There were no significant differences between the groups with respect to the prevalence of CAC score >100. Female athletes showed a lower prevalence of CAC score >50% centile (19% versus 32%; P=0.03) and CAC score >75% centile (14% versus 25%; P=0.045) compared with controls (**Figure 1**). Athletes were also less likely to exhibit pathological CAD (CAC score >100, or >75^th^ percentile for age) compared with controls (15% versus 25%; P=0.045).

**Figure 1.**
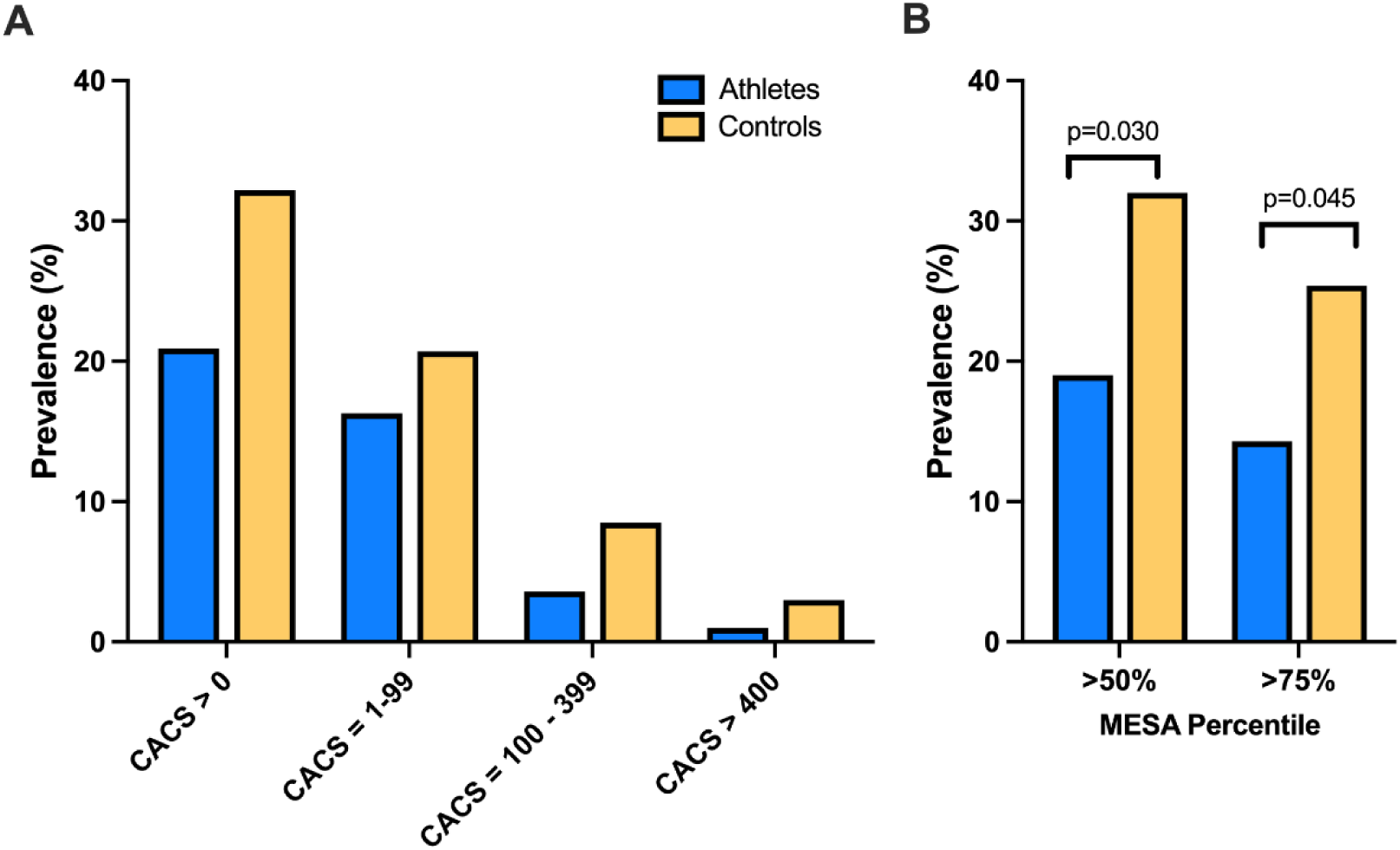
Coronary artery calcium (CAC) score characteristics in female master athletes and controls. The prevalence of CAC score categories did not differ between athletes versus controls (panel A), whereas athletes had a significantly lower prevalence of a CAC score exceeding the 50^th^ and 75^th^ Multi-Ethnic Study of Atherosclerosis (MESA) percentile (panel B).

**Table 2.** Comparison of CAC score, plaque prevalence, distribution and coronary inflammation in female master athletes and sedentary controls. Expressed as n (%)

|  | **Athletes**<br>**n=196** | **Controls**<br>**n=59** | **p-value** |
| --- | --- | --- | --- |
| Median CACS (AU) | 0 (0 - 0) | 0 (0 - 7) | 0.08 |
| CACS > 0 AU | 41 (21%) | 19 (32%) | 0.07 |
| CACS > 100 AU | 7 (3.6%) | 5 (8.5%) | 0.12 |
| CACS > 400 AU | 2 (1%) | 2 (3%) | 0.20 |
| CACS centile >50% | 37 (19%) | 19 (32%) | **0.03** |
| CACS centile >75% | 28 (14%) | 15 (25%) | **0.045** |
| Individuals with<br>≥1 Plaques | 42 (21%) | 19 (32%) | 0.09 |
| Individuals with solely<br>calcified plaques | 27 (14%) | 12 (20%) | 0.22 |
| Individuals with a<br>combination of calcified<br>and non-calcified plaques | 15 (8%) | 7 (12%) | 0.31 |
| Total number of plaques |  |  |  |
| Calcific | 61(80%) | 24 (63%) | 0.08 |
| Mixed morphology | 15 (18%) | 14 (37%) | **0.03** |
| Non calcific only | 1 (0.5%) | 0 | n/a |
| Left main plaque | 4 (2%) | 1 (2%) | 0.86 |
| Left anterior descending | 36 (16%) | 9 (15%) | 0.58 |
| Circumflex coronary artery | 6 (1%) | 3 (5.1%) | 0.46 |
| Right coronary artery | 11 (5.1%) | 4 (6.8%) | 0.74 |
| Number of vessels |  |  |  |
| 1-vessel disease | 32 (16%) | 13 (22%) | 0.20 |
| 2-vessel disease | 7 (4%) | 2 (3.4%) |  |
| 3-vessel disease | 3 (1.5%) | 4 (6.8%) |  |
| Stenosis >50% | 0 (0%) | 0 (0%) |  |
| Stenosis 25-49% | 6 (3%) | 4 (7%) | 0.22 |
| Stenosis <25% | 36 (18%) | 15 (25%) |  |

AU= Agatston Units; CACS= Coronary Artery Calcium Score; CAC = Coronary artery calcium; CAD = Coronary artery disease, FAI-RCA = Fat Attenuation Index in the Right Coronary Artery; LMCA = Left main coronary artery. Values are presented as n (percentage) or medians (Interquartile range 25%, 75%).

### Coronary plaques and luminal irregularities

Both athletes and controls had a low prevalence of atherosclerotic plaques (21% versus 32%; P=0.09). Athletes and controls did not differ in the prevalence or distribution of the atherosclerotic plaques (**Figure 2**, **Table 2**). Coronary plaques were predominantly located in the proximal and mid portion of the left anterior descending athletes among athletes and non-athletes. None of the female athletes or controls revealed obstructive CAD (luminal stenosis >50%), but 6 (3%) athletes and 4 (7%) controls had mild stenosis (25-49%) (**Table 2**). Coronary plaques were predominantly calcified in both athletes and controls (80% versus 63%; P=0.08) although athletes had a lower prevalence of mixed morphology plaques **(Figure 2b).**

**Figure 2.**
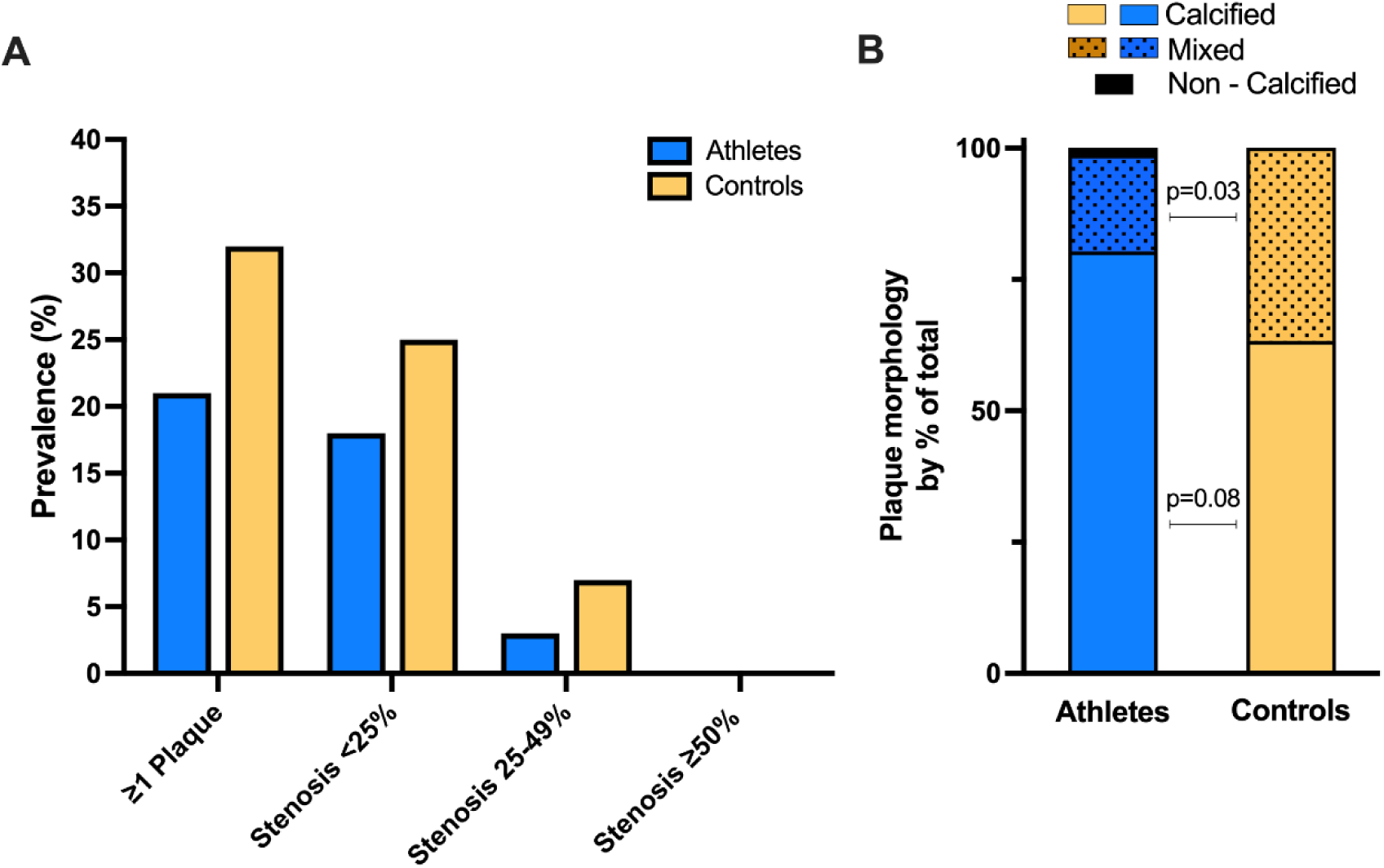
**A.** Presence of coronary atherosclerotic plaque and degree of stenosis in female master athletes and controls. **B.** Plaque morphology in female athletes (76 coronary plaques) and relatively sedentary males (38 coronary plaques).

### Coronary plaque volume, burden, and composition and coronary inflammation

Quantitative analysis revealed no differences in total, calcified, low density calcified, and non-calcified plaque volumes between the two groups (**Figure 3**, **Table 3**) In contrast, a higher burden of calcified plaque was observed in controls.

**Figure 3.**
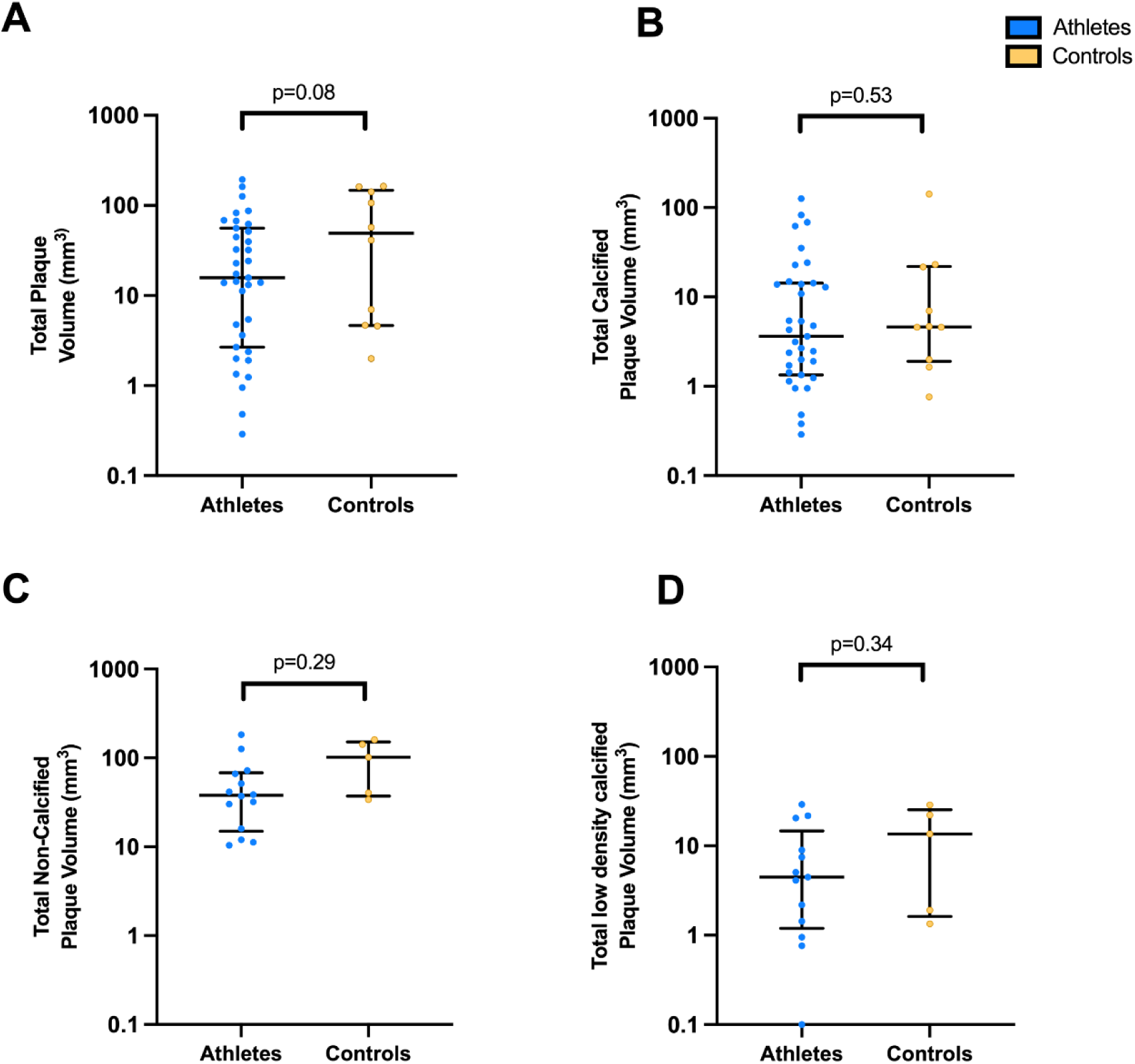
Quantitative characteristics of coronary atherosclerotic plaque in female master athletes and controls. Total plaque volume (A), calcified plaque volume (B), non-calcified plaque volume (C) and low-density calcified plaque volume (D) did not differ between athletes versus controls.

**Table 3.**
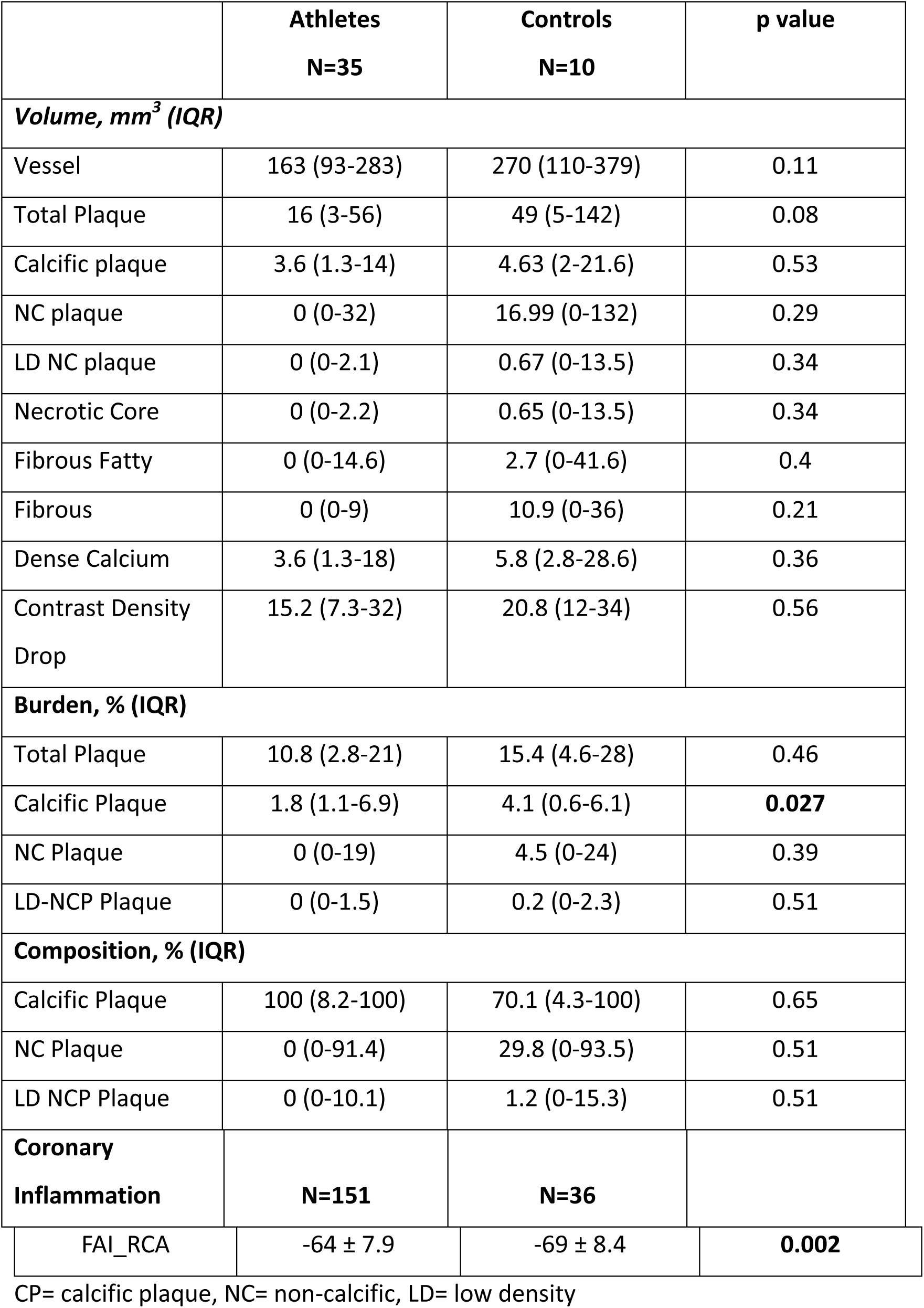
Comparison of plaque volume, burden and composition in master athletes and controls. Values are presented as median and 75%-25% interquartile range (IQR) if not otherwise indicated.

|  | **Athletes**<br>**N=35** | **Controls**<br>**N=10** | **p value** |
| --- | --- | --- | --- |
| **Volume, mm<sup>3</sup> (IQR)** |  |  |  |
| Vessel | 163 (93-283) | 270 (110-379) | 0.11 |
| Total Plaque | 16 (3-56) | 49 (5-142) | 0.08 |
| Calcific plaque | 3.6 (1.3-14) | 4.63 (2-21.6) | 0.53 |
| NC plaque | 0 (0-32) | 16.99 (0-132) | 0.29 |
| LD NC plaque | 0 (0-2.1) | 0.67 (0-13.5) | 0.34 |
| Necrotic Core | 0 (0-2.2) | 0.65 (0-13.5) | 0.34 |
| Fibrous Fatty | 0 (0-14.6) | 2.7 (0-41.6) | 0.4 |
| Fibrous | 0 (0-9) | 10.9 (0-36) | 0.21 |
| Dense Calcium | 3.6 (1.3-18) | 5.8 (2.8-28.6) | 0.36 |
| Contrast Density Drop | 15.2 (7.3-32) | 20.8 (12-34) | 0.56 |
| **Burden, % (IQR)** |  |  |  |
| Total Plaque | 10.8 (2.8-21) | 15.4 (4.6-28) | 0.46 |
| Calcific Plaque | 1.8 (1.1-6.9) | 4.1 (0.6-6.1) | **0.027** |
| NC Plaque | 0 (0-19) | 4.5 (0-24) | 0.39 |
| LD-NCP Plaque | 0 (0-1.5) | 0.2 (0-2.3) | 0.51 |
| **Composition, % (IQR)** |  |  |  |
| Calcific Plaque | 100 (8.2-100) | 70.1 (4.3-100) | 0.65 |
| NC Plaque | 0 (0-91.4) | 29.8 (0-93.5) | 0.51 |
| LD NCP Plaque | 0 (0-10.1) | 1.2 (0-15.3) | 0.51 |
| **Coronary Inflammation** | **N=151** | **N=36** |  |
| FAI_RCA | -64 ± 7.9 | -69 ± 8.4 | **0.002** |

CP= calcific plaque, NC= non-calcific, LD= low density

The level of coronary inflammation as derived by FAI_RCA_ was significantly higher (less negative) in female athletes compared with controls (-64 ± 7.9 HU versus -69 ± 8.4 HU, P=0.002 **(Table 3**; **Figure 4a).** Subgroup analysis revealed that FAI_RCA_ was significantly higher (more inflammation) in athletes with CAC score =0 compared to those with a CAC score >0) (-62.9± 7.3 versus -66.0 ± 8.8 HU; P=0.024) and compared to controls with a CAC score=0 (-62.9± 7.3 versus -68.2 ± 8.1 HU; P=0.001) (**Figure 4b**).

**Figure 4.**
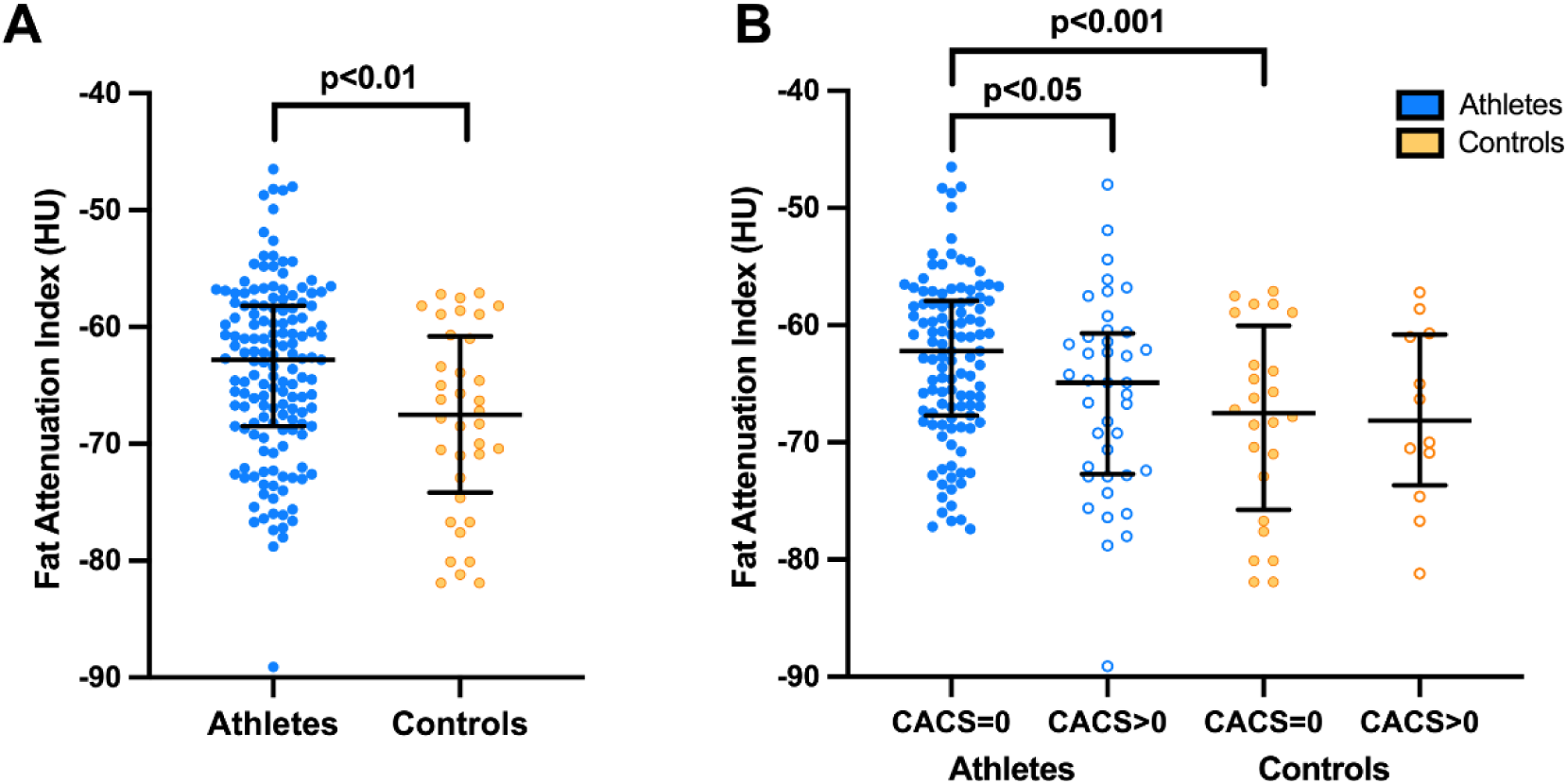
Fat attenuation index (FAI) measured in female master athletes and controls. Female athletes showed a significantly higher degree of coronary inflammation than controls (panel A). Athletes with a zero coronary artery calcium score **(CACS)** showed a higher FAI than athletes with **CACS**>0 (Panel B).

### Determinants of coronary atherosclerosis

Univariate analysis revealed that age, years of exercise, absolute peak VO2, resting systolic blood pressure, lower coronary inflammation (FAI_RCA_) and lower values of HDL-C, were associated with an increased probability of a CAC score>0. Multivariate logistic regression showed a positive association between age and CAC score> 0 (OR: 1.14 per year; 95% CI: 1.07–1.21); p<0.001] and resting SBP (OR: 1.03 per mmHg; 95% CI: 1.008–1.06); p=0.012] and an inverse association with HDL-C (OR: 0.34 per mol/L, 95% CI: 0.15-0.79; p=0.011) (**Table 4**). Among athletes, markers of coronary atherosclerosis were found in most women aged >65 years old (n=18) with 78% showing evidence of coronary plaque(s), 22% demonstrating a CAC score >100 AU and 11% revealing a CAC score >400 AU. Indeed, a CAC score >100 AU was almost confined to women aged ≥65 years old (**Figure 5**).

**Figure 5.**
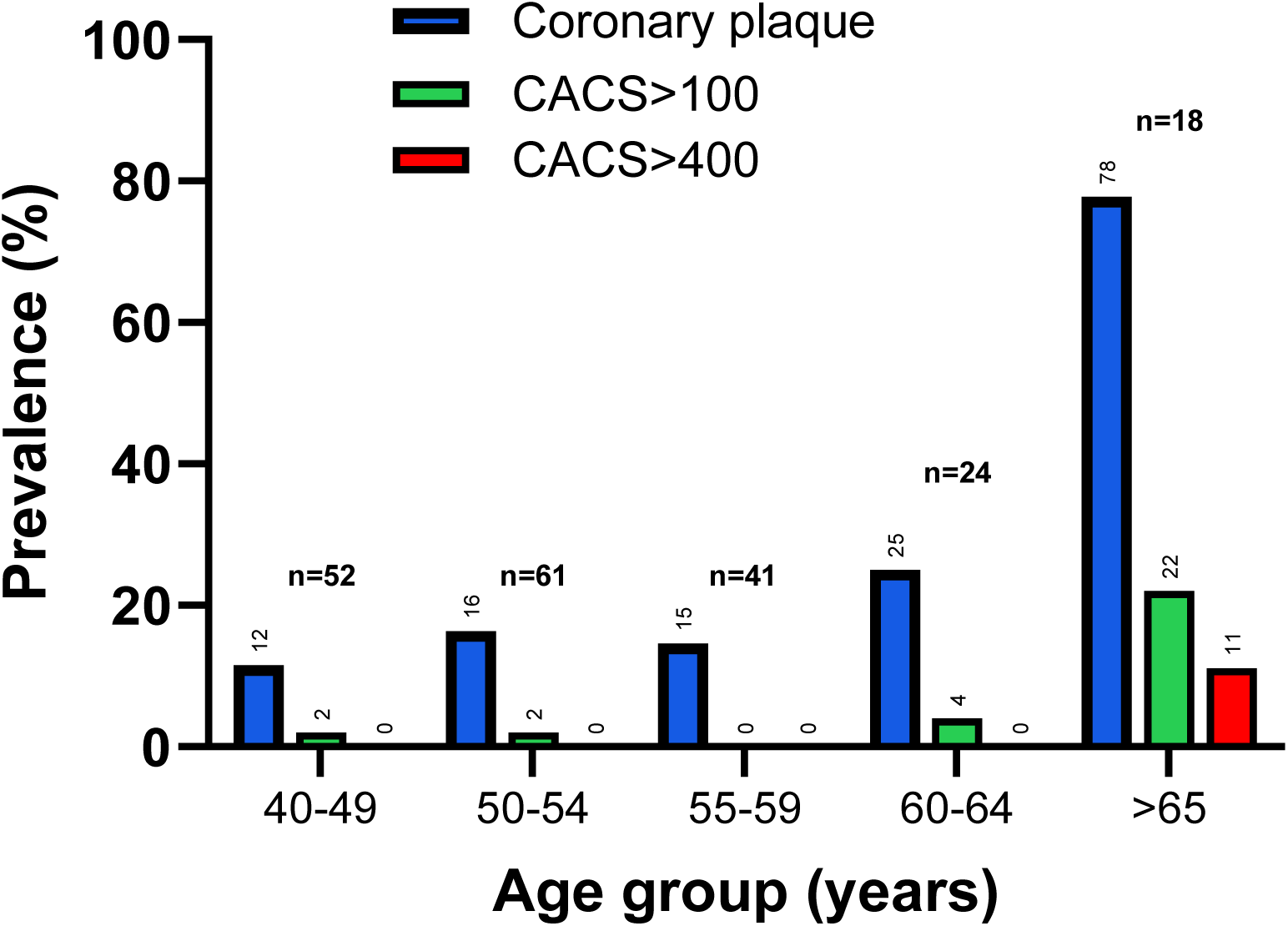
Coronary atherosclerosis in relation to age in female master athletes per different age groups. The presence of coronary artery calcium score (CACS) >100 AU and >400 AU was almost confined to female master athletes > 65 years old.

**Table 4.** Determinants of coronary atherosclerosis in female master athletes. Univariate and multivariate analysis.

|  | **CACS=0**<br>**N=155** | **CACS>0**<br>**N=41** | **Univariate** | **Multivariate**<br>**R<sup>2</sup>=0.21** |
| --- | --- | --- | --- | --- |
| Age | 53 ± 0.6 | 59.7 ± 1.5 | **<0.001** | **<0.001** |
| Menopausal | 107 (69%) | 32 (78%) | 0.29 |  |
| BMI (kg/m<sup>2</sup>) | 21.3 (20-22.6) | 21.6 (20-23.4) | 0.62 |  |
| Years of Exercise | 31 ± 11 | 35 ± 12 | **0.047** |  |
| Hours per Week | 8 (7-10) | 8 (7-11) | 0.93 |  |
| Days per Week | 6 (5-6) | 5.3 (4-6) | 0.24 |  |
| Total Competitions | 76 (30-185) | 100 (50-145) | 0.66 |  |
| Total METS min/w | 7239 (4784-10932) | 6835 (4743-9498) | 0.59 |  |
| Vigorous METS min/w | 2880 (2880-4800) | 2880 (2160-4800) | 0.92 |  |
| PTH levels (pmol/L) | 7.3 (6-9.4) | 7.7 (5.9-9.8) | 0.66 |  |
| Total cholesterol | 5.1 (4.7 – 6) | 5.4 (4.7 – 5.9) | 0.50 |  |
| HDL-C (mmol/L) | 2.5 ± 0.8 | 2.1 ± 0.7 | **0.007** | **0.011** |
| Peak VO<sub>2</sub> (ml/min/kg) | 41.6 ± 6.18 | 38.8 ± 8.35 | **0.020** |  |
| Peak VO<sub>2</sub> predicted (%) | 163 ± 23.4 | 165.5 ± 28.4 | 0.90 |  |
| FAI<sub>RCA</sub> (HU) | -63.6 ± 7.3 | -66.7 ± 8.8 | **0.024** |  |
| Resting SBP (mmHg) | 113 ± 14 | 122 ± 16 | **<0.001** | **0.012** |
| SBP>190 mmHg<sup>#</sup> | 36 (23%) | 13 (32%) | 0.28 |  |

FAI<sub>RCA</sub> = Fat Attenuation Index in the Right Coronary Artery; HDL-C= High Density Lipoprotein Cholesterol; HU = Hounsfield Units; METS = Metabolic Equivalents; PTH = Parathyroid Hormone; VO<sub>2</sub> = Oxygen Consumption, SBP: Systolic blood pressure.

### Determinants of coronary inflammation

A higher degree of coronary inflammation was associated with, years of exercise, peak VO2, a lower BMI and lower age and a higher HDL. Multivariate linear regression identified a positive correlation (R^2^=0.19) between inflammation and peak VO_2_ (beta 0.44 per ml/min/kg 95% CI 0.24-0.64; p<0.01) and HDL (beta 1.4 per mmol/L 95% CI 0.10-2.7; p<0.05).

### Myocardial scar

Five athletes had major scar within the left ventricular myocardium including 2 with an ischaemic pattern (one with a small area of subendocardial basal inferolateral scar and the other with a small area of focal transmural basal inferior scar) and 3 athletes with non-ischaemic scar affecting the mid portion of the lateral LV wall. Neither of the 2 athletes with ischaemic scar revealed any evidence of CAD on CCTA.

## DISCUSSION

There are few existing studies reporting coronary atherosclerosis in master female athletes with a cumulative total of <100 athletes^6,10–12^. Based on the relatively small numbers and contradictory conclusions in these small studies, it is difficult to infer whether increased CAC and plaque burden observed in some female athletes reflects advanced age, cumulative effects of prodigious exercise, or other factors.

In the current study, we investigated the prevalence of markers of coronary atherosclerosis in 196 female master athletes and 59 ostensibly healthy and relatively sedentary women who were predominantly post-menopausal and had a low atherosclerotic risk profile. Despite a significantly larger cohort of athletes, we did not observe any differences in CAC score, number of coronary plaques, plaque volume or the morphology of the coronary plaques between the two groups. Both athletes (21%) and controls (32%) had a low prevalence of CAD, although athletes were less likely to have a CAC score >50^th^ and 75^th^ percentile for age. Age was the most important determinant of an elevated CAC score and most females with markers of coronary atherosclerosis were aged ≥65years old.

### Predictors of coronary atherosclerosis

Exercise duration and volume have been associated with CCTA markers of atherosclerosis in male athletes^3^, however we did not observe any significant association with the exercise history and CAC score even though our female athletes had exercised for a median of 33 years and participated in a median of 90 endurance events, which is comparable with previous studies in male athletes with raised CAC score^3,4,6^. Several mechanisms have been implicated for the increased coronary atherosclerosis in male master athletes^7,8,22^, including damage from mechanical stresses of bending and flexing of the coronary arteries, increased LV mass, exercise-induced hypertension^23^, post-exertional increased parathyroid levels^24,25^ and coronary inflammation^26^. We observed a SBP>190mmHg during exercise in a quarter of our athletes, but could not reveal any association with CAD. Similarly, exercise-induced elevations of PTH concentrations were observed, but the magnitude of these changes were equal in athletes and controls and did not correlate with coronary calcification. In contrast, traditional risk factors such as age, resting blood pressure and lipid profile influenced CAC score in female athletes. As opposed to previous studies in master male athletes, we could not demonstrate any association between the duration and volume of exercise and an elevated CAC ^3,27^.

### Coronary Inflammation

Several studies have indicated that there may be a threshold beyond which high-intensity endurance exercise can cause increased oxidative stress and reduced levels of anti-oxidants^28,29^. Schwarz *et al* revealed an acute and transient increase in post-race apoptotic endothelial microparticles in 99 middle-aged marathon runners, including 22% females, indicating that high intensity exercise may be linked to endothelial cell damage and a pro-inflammatory response^30^. Lin *et al*^26^ investigated coronary artery plaque characteristics before and after completion of a 140-day race entailing 25.7 miles/day with one rest day per week. Among the 4 runners with prior CAD, there was an increase in non-calcified plaque volume at sites of pre-existing disease, whereas there was no evidence of *de novo* plaque in the remaining 5 runners. These observations suggest that extreme endurance exercise may stimulate CAD progression independent of traditional atherosclerotic factors, through mechanisms including inflammation. We revealed an increased prevalence of coronary inflammation in our female athletes compared with controls, however this was not a determinant or associated with a raised CAC score. Conversely, there appeared to be an inverse trend between inflammation in athletes with a normal CAC score versus athletes with an abnormal CAC score. It should be noted, however, that most female athletes in our cohort had a zero CAC score and individuals with conventional risk markers for endothelial damage had been excluded from the study.

### Sex disparities

Our data suggest that there is sex-specific coronary artery remodeling in master athletes. Whereas our previous experience in male athletes has shown a higher prevalence of high CAC (>100 AU) and increased prevalence of coronary plaques compared with healthy controls ^7–9^ we did not observe similar results in our female master athletes. In contrast, female master athletes were less likely to have a CAC>50^th^ and 75^th^ centile and mixed morphology plaques compared with healthy non athletes. Age was the most important determinant of CAD. Although older female athletes (>65-years-old) showed a higher prevalence of CAC score>0 and plaque burden compared with younger women, we observed exclusively non-obstructive CAD in all age categories compared with previous reports of significant stenoses (>50%) in 6-8% males^6,27^. Sex-specific characteristics including hormonal ^31,32^, hemodynamic, anthropometric, and psychological factors may be protective in female master athletes compared with male athletes.

Despite the lack of correlation between previously speculated associations between exercise and markers of coronary atherosclerosis on CCTA among our female athletes, we identified ischaemic scar in two athletes who had no evidence of CAD, suggesting that there may be other mechanisms for limited infarction, such as coronary spasm, demand ischaemia or spontaneous occlusion of micro vessels due to a prothrombotic state although our numbers are too small to make any strong inferences.

### Limitations

Our study has several limitations. We relied solely on subjective assessment for menstrual status rather than endocrine profiles, therefore we cannot be certain how far our results were affected by circulating estrogen concentrations. We did not investigate the effect of dietary habits or genetic predisposition in the development of atherosclerosis in our athletes. Most of our athletes were white, and all were involved in endurance sports therefore it is uncertain whether these results can be extrapolated in athletes of different ethnicity or sports disciplines.

## CONCLUSION

In contrast with previous experience in male master endurance athletes, female counterparts do not reveal increased CAC scores or plaque burden compared with relatively sedentary healthy women. Among the small number of female athletes with increased CAC scores, age, blood pressure and lipid profile were the most important determinants of coronary atherosclerosis rather than the volume, duration and intensity of exercise. Larger studies in even older master female athletes may help delineate sex related differences in coronary artery remodeling in lifelong endurance athletes more precisely.

## Data Availability

Data available on request from the authors

## Acknowledgements

The authors would like to acknowledge 1. Dr Sarojini David for assisting and interpreting the coronary computed tomography angiograms. 2. Paulo Bulleros and Zephryn Fanton for assistance with transthoracic echocardiography and cardiopulmonary exercise tests.

## Funding

Dr Efstathios Papatheodorou was funded by a research grant provided by the charitable organisation Cardiac Risk in the Young (CRY).

## Disclosures

None of the authors have any conflicts to declare.

## Notes

### Competing Interest Statement

The authors have declared no competing interest.

### Author Declarations

NHS Research Ethics Committee reference: 13/SW/0163

